# Self-Reported Stress and Questionnaires in People With Autism Spectrum Disorder: a Systematic Review

**DOI:** 10.1101/2021.02.27.21252281

**Authors:** Anoushka Thoen, Jean Steyaert, Kaat Alaerts, Kris Evers, Tine Van Damme

## Abstract

**Background:** To gain more insight into the experience of stress in individuals with ASD, it is important to use appropriate self-report questionnaires. The goal of this systematic review was to provide an overview of these.

**Method:** The PRISMA guidelines were followed and four online databases were systematically searched.

**Results:** Seven questionnaires have been used previously in individuals with ASD. None of the 22 included studies intended to assess the psychometric properties, leading towards scarce evidence concerning their reliability and validity in this population.

**Conclusions:** It is important to consider which concept of stress one aims to measure as not all questionnaires cover the same aspects of stress. Further research concerning psychometric properties of the questionnaires in this population is required.

---

Children and adults with Autism Spectrum Disorder (ASD) report higher levels of stress compared to typically developing individuals (Bishop-Fitzpatrick et al., 2015, 2017a; Browning et al., 2009; Groden, 2006; Hirvikoski & Blomqvist, 2015; McGillivray & Evert, 2018). A reciprocal relationship between the severity of ASD-symptoms and high levels of perceived stress has been found in previous research (Bishop□Fitzpatrick et al., 2015; Groden, 2006; Hirvikoski & Blomqvist, 2015; Pahnke et al., 2014; Porges et al., 2013). Additionally, ASD-symptoms may also restrict the ability of these individuals to seek for help or social support when needed (Hirvikoski & Blomqvist, 2015). This may lead to long-term presence of high levels of stress, which has a profound effect on physical and mental health, as demonstrated in typically developing individuals (Mendelson, 2013; Slavich, 2016).

The most frequently reported information in stress research covers objective features related to the stress response, including changes at physiological and behavioral levels. Yet, another important aspect of stress is the level of perceived stress, which is defined as ‘the feelings and thoughts an individual has related to the stressfulness of their life and their ability to overcome stressful events’ (Phillips, 2013). As these thoughts and feelings are related to factors such as personality, coping resources and support, individuals may encounter similar negative life events but can appraise the impact or severity differently. This aspect could be referred to as the subjective information concerning stress and should rely on self-reported measures. In several studies, Bishop et al. (2015, 2017a, 2018) have reported that in adults with ASD, with and without co-occurring intellectual disability, high levels of perceived stress were associated with poor social functioning, social outcome and quality of life. In addition, it has been stated that interventions for adults with ASD may be less efficient due to the high levels of perceived stress as these may hamper the use of learned cognitive strategies to control behavior (Bishop-Fitzpatrick et al., 2017a). These findings point towards the clinical significance of perceived stress and its assessment in individuals with ASD. However, contrary to research involving the stress response, little research attention has been paid to this subjective component in individuals with ASD. This underrepresentation of research may relate to the fact that individuals with ASD often display difficulties with reporting on their own affective states (DuBois et al., 2016). In addition, symptoms of stress in individuals with ASD may have been coupled to other concepts such as quality of life, mood symptoms and problems with emotion regulation (Bishop-Fitzpatrick et al., 2017a). Accordingly, self-reporting tools have often been perceived as inaccurate and unreliable in individuals with ASD (Baron et al., 2006), leading to a scarcity of information regarding, for instance, perceived stress in this population. However, in recent years the field is evolving, acknowledging the increased need for reliable self-report tools in individuals with ASD. Recent work in the field of internalizing states has provided promising evidence regarding the reliability and validity of self-report tools (Berthoz & Hill, 2005; Keith et al., 2019; Rieffe et al., 2011), and a similar tendency is seen for self-reports on stress.

Therefore, this systematic review provides an exhaustive overview of self-report measures of stress in individuals with ASD, including a description of their construct and psychometric properties, where available. In addition, some studies might not use the specific term of perceived stress but instead refer only to the measurement of self-reported stress. Therefore, the compliance towards the definition of perceived stress as described by Phillips (2013) will be discussed as well.

## Methods

This systematic review was performed according to the Preferred Reporting Items for Systematic Review and Meta-Analysis (PRISMA) statement (see electronic supplementary material ESM-1). Analysis of methodological quality, which is obligatory following the PRISMA-guidelines, was not feasible for the included studies, as further explained in the discussion.

### Information Sources and Search Strategy

The search strategy, based on the Population Intervention Comparison Outcome (PICO) method, was entered in PubMed, Embase, Web of Science and Cochrane Library in November 2019 and was last updated in May 2020. A combination of free text words, controlled terminology (f.i. MeSH-terms) and linguistic variations was used based on the concepts of ‘stress’ and ‘Autism Spectrum Disorder’ (see electronic supplementary material ESM-2). No filters were applied.

### Eligibility Criteria and Screening Procedure

In order to be included, studies needed to fulfil the following eligibility criteria: (1) a study population older than 6 years; (2) an ASD-diagnosis according to the Diagnostic and Statistical Manual of Mental Disorders (American Psychiatric Association, 1980, 1987, 1994, 2000, 2013) or the International Statistical Classification of Diseases and Related Health Problems (World Health Organization, 2016, 2019); (3) use of self-reported questionnaires as a stress assessment tool; (4) original research written in English or Dutch. Reviews, meta-analyses, qualitative designs, case studies/-series, editorials, conference papers, books and book chapters, trial registrations, letters to the editor, abstracts only and expert opinions were excluded.

The selection process consisted of two phases. The first phase was conducted by one researcher (AT), who screened the title and abstract following the eligibility criteria as mentioned above. Articles were included if they met these criteria or in case eligibility could not yet be determined. In the second phase, two independent researchers (AT and TVD) screened the full texts of these articles following the same eligibility criteria. During a consensus meeting, any doubts or disagreements were discussed. In addition, the reference lists of the included studies were screened and additional articles were included if eligible.

### Data Extraction and Questionnaire Evaluation

Two researchers (AT and TVD) performed data extraction based on the following variables: population characteristics of individuals with ASD (diagnosis, age, gender and exclusion criteria), the employed questionnaire and information concerning the content and construct of the questionnaire. Information regarding psychometric properties of the questionnaires was not requested but if present, related data was extracted from the studies (Tables 1 and 2). The following items concerning psychometric properties were gathered: internal consistency, reliability and validity measures and the presence of data from a comparison group within the same study. The latter item was included to provide preliminary evidence of construct validity, confirming the hypothesis that higher scores were reported in individuals with ASD in comparison to control groups. Any disagreements or doubts were discussed during a consensus meeting. Information concerning psychometric properties, determined in other populations, fall outside the scope of this review but have been described in previous reports for most of the included questionnaires.

**Table 1.**
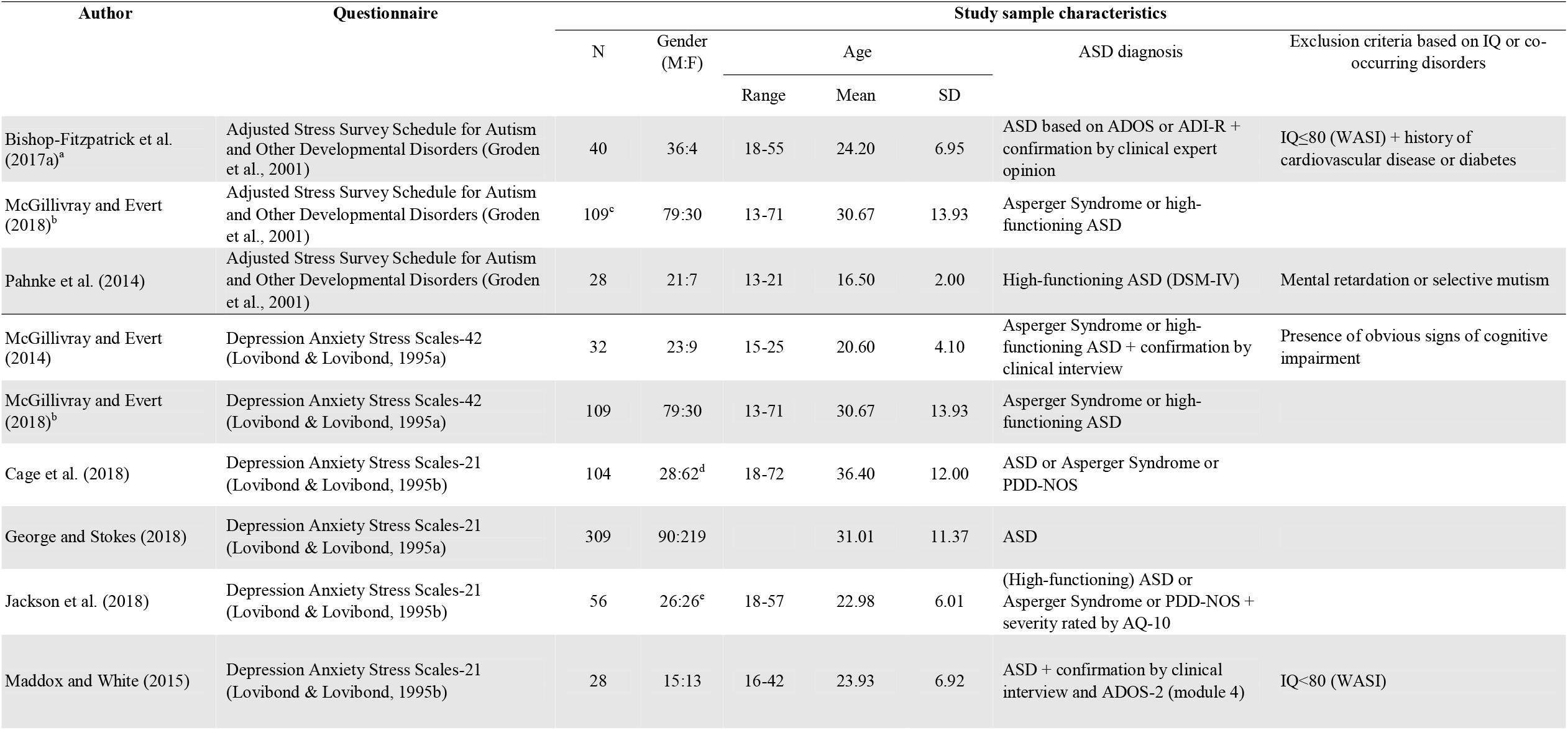
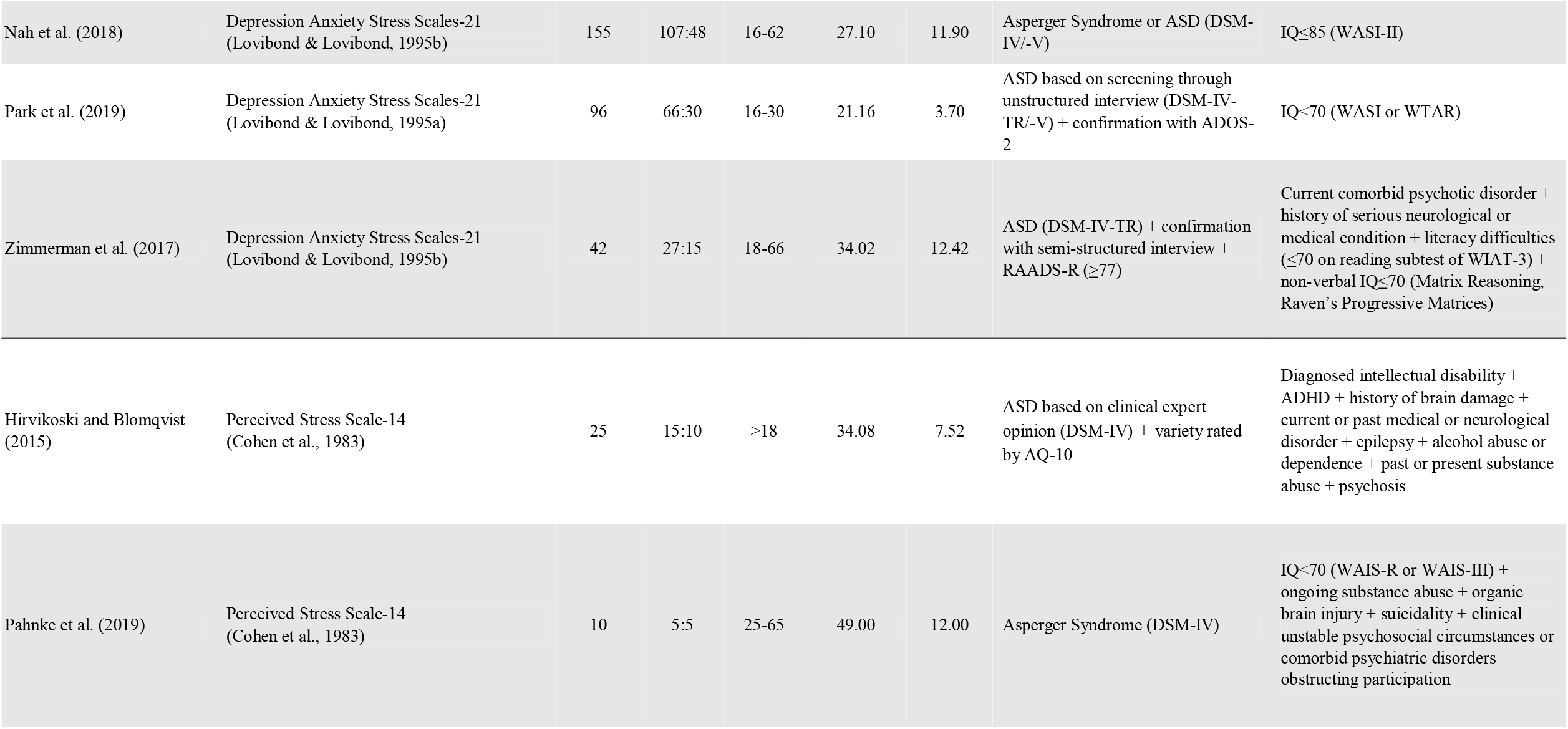
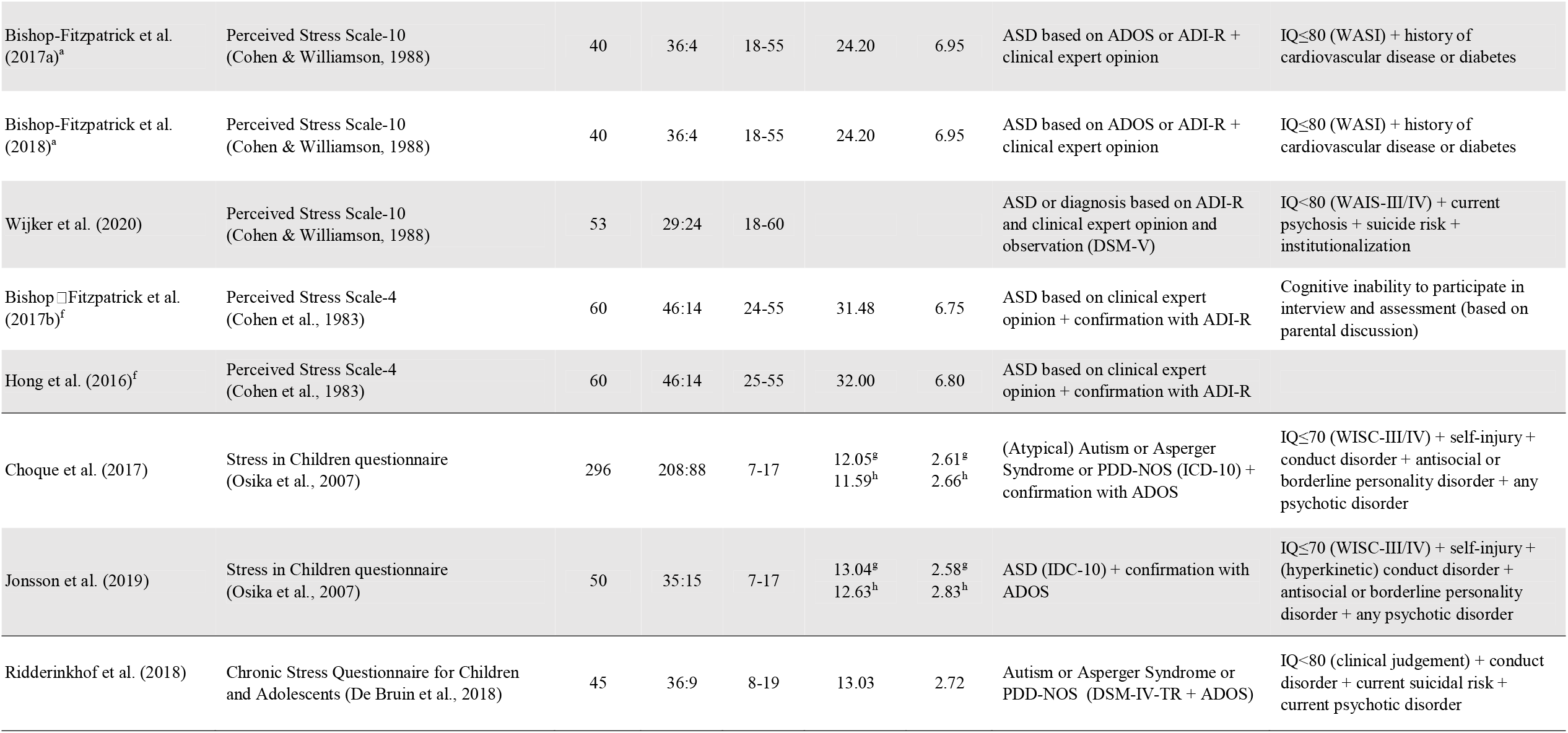
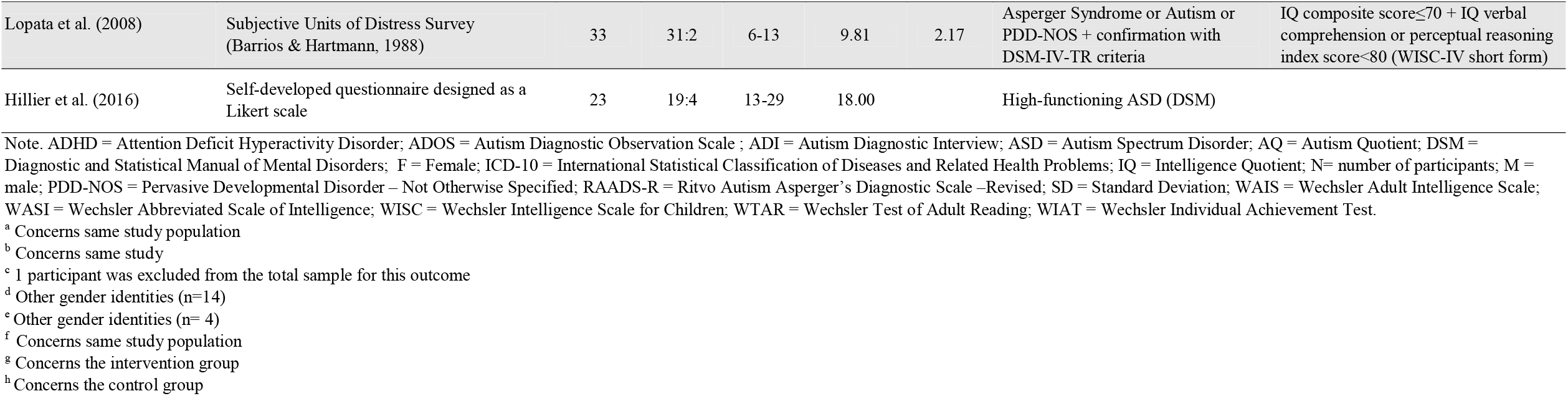
Characteristics of the Individual Studies

**Table 2.**
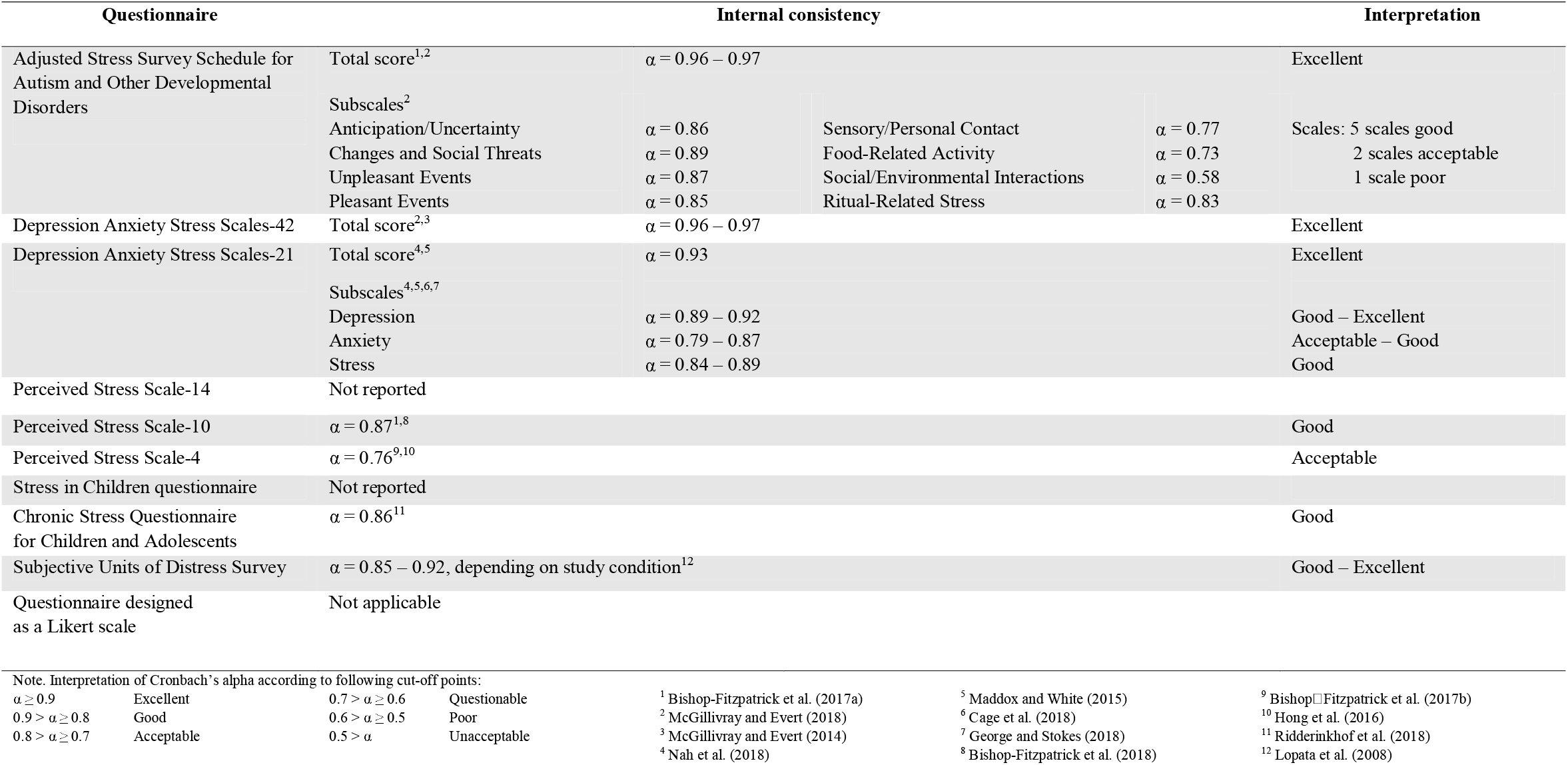
Overview of Internal Consistency of Included Questionnaires

| Questionnaire |  | Internal consistency |  |  | Interpretation |
| --- | --- | --- | --- | --- | --- |
| Adjusted Stress Survey Schedule for Autism and Other Developmental Disorders | Total score<sup>1,2</sup> | $\alpha = 0.96 - 0.97$ | | | Excellent |
|  | Subscales<sup>2</sup> |  |  |  |  |
| | Anticipation/Uncertainty | $\alpha = 0.86$ | Sensory/Personal Contact | $\alpha = 0.77$ | Scales: 5 scales good<br>2 scales acceptable<br>1 scale poor |
| | Changes and Social Threats | $\alpha = 0.89$ | Food-Related Activity | $\alpha = 0.73$ | |
| | Unpleasant Events | $\alpha = 0.87$ | Social/Environmental Interactions | $\alpha = 0.58$ | |
| | Pleasant Events | $\alpha = 0.85$ | Ritual-Related Stress | $\alpha = 0.83$ | |
| Depression Anxiety Stress Scales-42 | Total score<sup>2,3</sup> | $\alpha = 0.96 - 0.97$ | | | Excellent |
| Depression Anxiety Stress Scales-21 | Total score<sup>4,5</sup> | $\alpha = 0.93$ | | | Excellent |
|  | Subscales<sup>4,5,6,7</sup> |  |  |  |  |
| | Depression | $\alpha = 0.89 - 0.92$ | | | Good – Excellent |
| | Anxiety | $\alpha = 0.79 - 0.87$ | | | Acceptable – Good |
| | Stress | $\alpha = 0.84 - 0.89$ | | | Good |
| Perceived Stress Scale-14 | Not reported |  |  |  |  |
| Perceived Stress Scale-10 | $\alpha = 0.87^{1,8}$ | | | | Good |
| Perceived Stress Scale-4 | $\alpha = 0.76^{9,10}$ | | | | Acceptable |
| Stress in Children questionnaire | Not reported |  |  |  |  |
| Chronic Stress Questionnaire for Children and Adolescents | $\alpha = 0.86^{11}$ | | | | Good |
| Subjective Units of Distress Survey | $\alpha = 0.85 - 0.92$, depending on study condition<sup>12</sup> | | | | Good – Excellent |
| Questionnaire designed as a Likert scale | Not applicable |  |  |  |  |

Note. Interpretation of Cronbach's alpha according to following cut-off points:

|  |  |  |  |
| --- | --- | --- | --- |
| $\alpha \geq 0.9$ | Excellent | $0.7 > \alpha \geq 0.6$ | Questionable |
| $0.9 > \alpha \geq 0.8$ | Good | $0.6 > \alpha \geq 0.5$ | Poor |
| $0.8 > \alpha \geq 0.7$ | Acceptable | $0.5 > \alpha$ | Unacceptable |

<sup>1</sup> Bishop-Fitzpatrick et al. (2017a)
<sup>2</sup> McGillivray and Evert (2018)
<sup>3</sup> McGillivray and Evert (2014)
<sup>4</sup> Nah et al. (2018)
<sup>5</sup> Maddox and White (2015)
<sup>6</sup> Cage et al. (2018)
<sup>7</sup> George and Stokes (2018)
<sup>8</sup> Bishop-Fitzpatrick et al. (2018)
<sup>9</sup> Bishop-Fitzpatrick et al. (2017b)
<sup>10</sup> Hong et al. (2016)
<sup>11</sup> Ridderinkhof et al. (2018)
<sup>12</sup> Lopata et al. (2008)

For clarity, a categorization in *general ‘trait-like’ stress measures* and *moment-specific ‘state-like’ stress measures* was used. A trait is thereby considered as part of an individual’s personality, thus a long-term characteristic, whereas a state is influenced by external events, thus temporary (The Oxford Review Encyclopaedia of Terms, 2019). The general measures were further divided in (i) questionnaires solely including *stress-specific questions* and (ii) *combined questionnaires* including other psychological symptoms.

## Results

### Study Selection and Population Characteristics

The search strategy in the four different databases resulted in 8363 articles after deduplication. After two screening phases, 20 articles were retained and two additional articles were included after reference screening, resulting in 22 included articles (for the selection flowchart, see Figure 1).

**Figure 1.**
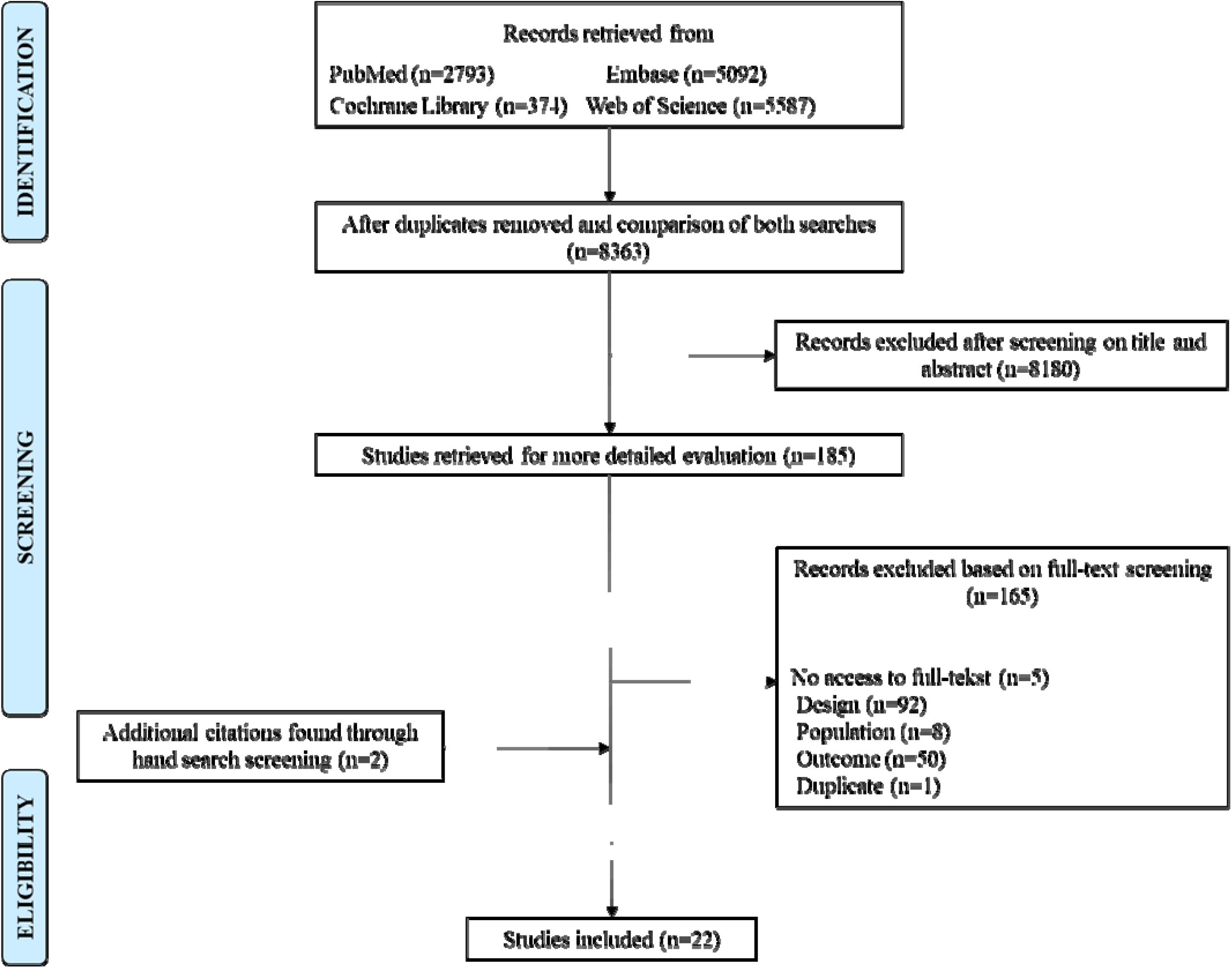
Flow-Chart of the Selection Process *[Figure created in Microsoft Office PowerPoint 2016]*

A total of 20 different study samples was identified, as two study samples were independently reported twice in four different articles (Table 1). In total, 1594 individuals with ASD were included from which 942 were male. The gender distribution represented a male preponderance in most study samples (in 16 out of 20) as typically found in populations with ASD (Giarelli et al., 2010), except for four studies reporting more females than males or an equal distribution between genders in their study sample. In addition, the ages ranged between 13 and 71 years with only four studies using self-reports of stress in children and adolescents (13-18 years). The most frequently used exclusion criteria were based on intellectual ability (Intelligence Quotient (IQ) ≤ 70, 80 or 85) and the presence of co-occurring psychiatric disorders or problems such as current psychotic disorders, suicide risk and substance abuse. The studies defined the presence of an ASD-diagnosis using different terminology including (high-functioning) ASD, Asperger’s Syndrome or PDD-NOS (Pervasive Developmental Disorder – Not Otherwise Specified). More detailed information can be found in Table 1. [Please insert tables at the end of the complete manuscript to enhance readability]

A total of seven different questionnaires was used to assess self-reported stress in individuals with ASD. More detailed information concerning the content and psychometric properties of the individual questionnaires is available in Table 2.

### General ‘Trait-Like’ Stress Questionnaires

#### Stress-Specific Questionnaires

This category is characterized by questionnaires focusing on the frequency of behavior, feelings and/or somatic problems related to stress or on the intensity of the stress reaction. Five questionnaires fulfilled this description: (i) the Adjusted Stress Survey Schedule (SSS; Groden et al., 2001), (ii) the Perceived Stress Scale (PSS; Cohen et al., 1983; Cohen & Williamson, 1988), (iii) the Stress in Children Questionnaire (SiC questionnaire; Osika et al., 2007), (iv) the Chronic Stress Questionnaire for Children and Adolescents (CSQ-CA; De Bruin et al., 2018) and (v) a self-developed questionnaire (Hillier et al., 2016).

The *SSS* was the only questionnaire included in this review that was specifically developed for individuals with ASD (Groden et al., 2001). Although its original version constituted an informant-report measure, a modified self-report version of the SSS for adolescents and adults with ASD was recently developed and used in three studies (Bishop-Fitzpatrick et al., 2017a; McGillivray & Evert, 2018; Pahnke et al., 2014). The respondents are asked to rate the intensity of their stress reaction in common daily activities, divided into eight categories: (i) Changes and Threats, (ii) Anticipation/Uncertainty, (iii) Unpleasant Events, (iv) Pleasant Events (such as presents or birthday parties), (v) Sensory/Personal Contact, (vi) Food-Related Activity, (vii) Social/Environment Interactions and (viii) Ritual-Related Stress (Groden et al., 2001). In two studies, Cronbach’s alpha was reported (Bishop-Fitzpatrick et al., 2017a; McGillivray & Evert, 2018), indicating excellent internal consistency of the questionnaires’ total score (α = 0.96-0.97). The internal consistency of the subscales, ranging from 0.58 to 0.89, reported in the study of McGillivray and Evert (2018) was similar to the internal consistency values of the original version by Groden et al. (2001). One study compared the scores on the SSS of adults with ASD to those of typical peers and found statistically significant differences, indicating a higher stress intensity for adults with ASD (Bishop-Fitzpatrick et al., 2017a).

The *PSS* was developed as a self-report questionnaire and designed to measure “the degree to which individuals appraise situations in their lives as stressful” (Cohen et al., 1983). The items focus on stress-related behaviors and feelings. Three versions of the PSS exist, with the original version containing 14 items (PSS-14), followed by the development of two shorter versions that contain 10 (PSS-10) and 4 (PSS-4) items (Cohen & Williamson, 1988). In two studies, the original PSS-14 was used in adults with ASD, but information on psychometric properties was not reported (Hirvikoski & Blomqvist, 2015; Pahnke et al., 2019). The PSS-10 was used in three studies using two different adult populations with ASD (Bishop-Fitzpatrick et al., 2017a, 2018; Wijker et al., 2020). Two studies with the same study sample measured the degree of perceived stress during the last month (Bishop-Fitzpatrick et al., 2017a, 2018) while the third study did not specify the examined timespan (Wijker et al., 2020). Good internal consistency was reported in two studies with a Cronbach’s alpha of 0.87 (Bishop-Fitzpatrick et al., 2017a, 2018). Lastly, the PSS-4 was reported in two studies using the same adult population with ASD (Bishop□Fitzpatrick et al., 2017b; Hong et al., 2016). Both studies specified the examined timespan as ‘during the last month’ and reported an acceptable internal consistency, as rated by Cronbach’s alpha (α = 0.76). Only one study compared the scores on the PSS-14 of adults with ASD to those of typical peers (Hirvikoski & Blomqvist, 2015). Results indicated that adults with ASD reported significantly higher perceived stress regarding the PSS-14 total score and both subscales.

The *SiC questionnaire* was developed as a self-report questionnaire by Osika and colleagues to assess the degree of perceived distress in children (Osika et al., 2007). In addition, the presence of symptoms of lower well-being and important aspects of coping and social support are examined as well. Children need to rate the frequency of 21 physical and emotional symptoms of stress on a 4-point Likert scale. Two studies have used this questionnaire in children and adolescents with ASD (7-17 years; Choque et al., 2017; Jonsson et al., 2019). The developers of this questionnaire have advised the use of cut-off criteria to categorize the children’s stress level as following: ‘No stress’ (<2), ‘Medium stress’ (2-2.5) and ‘High stress’ (≥2.5) (Stallknecht et al., 2017). However, these were not applied in the two studies included in this review (Choque et al., 2017; Jonsson et al., 2019). In addition, measures of psychometric properties were not reported.

The *CSQ-CA* was used in one study in children and adolescents with ASD to rate the personal relevance of 19 described feelings and behaviors during the past 3 months. Internal consistency was good, as rated by Cronbach’s alpha (α = 0.86; Ridderinkhof et al., 2018).

One study used a *self-developed questionnaire* consisting of a Likert scale to measure the degree of perceived stress on an average day in adolescents and young adults with ASD (Hillier et al., 2016). Information concerning the scaling or psychometric properties was not reported.

#### Combined questionnaires

This category contains only one questionnaire, the Depression Anxiety Stress Scale (DASS; Lovibond & Lovibond, 1995a). The *DASS* contains items reflecting on symptoms of depression, stress and anxiety. Respondents are asked to rate the frequency of these symptoms during the past week. Two studies used the original 42-item version in adolescent and adult populations with ASD and reported excellent internal consistency of the scales’ total score (α = 0.96-0.97; McGillivray & Evert, 2014, 2018). In addition, one study reported higher mean scores on all subscales in young adults with ASD (McGillivray & Evert, 2018) compared to the normative data from the DASS manual (Lovibond & Lovibond, 1995b). Seven studies used the short 21-item version in adolescents and adults with ASD from which three studies specified the timespan examined as ‘during the past week’, following the manual’s instructions (Cage et al., 2018; Maddox & White, 2015; Zimmerman et al., 2017). One study used the DASS-21 to measure current levels of symptoms (Jackson et al., 2018) whereas the remaining three studies did not specify in which timespan the symptoms had to be present (George & Stokes, 2018; Nah et al., 2018; Park et al., 2019). Measures of internal consistency were reported in five studies, with a Cronbach’s alpha of 0.93 for the scales’ total scores (Maddox & White, 2015; Nah et al., 2018), 0.89 to 0.92 for the depression subscale, 0.79 to 0.87 for the anxiety subscale and 0.84 to 0.89 for the stress subscale score (Cage et al., 2018; George & Stokes, 2018; Maddox & White, 2015; Nah et al., 2018). Five studies using the DASS-21 compared data from a study sample of individuals with ASD with data from typical peers or norm values, a clinical population or another population with ASD and co-occurring social anxiety disorder. Individuals with ASD scored significantly higher on all subscales (Cage et al., 2018; George & Stokes, 2018; Nah et al., 2018) and the total score (Maddox & White, 2015; Nah et al., 2018) compared to typical peers or norm values. One study reported significantly lower subscale scores in individuals with ASD in comparison to a clinical group with anxiety and depression (Nah et al., 2018). Another study reported significantly higher subscale scores in young adults with ASD but only in comparison to young adults with psychosis and not in comparison to young adults with depression, anxiety or bipolar disorder (Park et al., 2019). A clinical comparison group with social anxiety disorder did not differ significantly on the DASS total score in comparison to individuals with ASD (Maddox & White, 2015). In contrast, individuals with ASD and co-occurring social anxiety disorder did score significantly higher on the DASS total score and the stress subscale when compared to individuals with ASD without co-occurring social anxiety disorder (Maddox & White, 2015).

### Moment-Specific ‘State-Like’ Questionnaire

Only the Subjective Units of Distress Survey (SUDS; Barrios & Hartmann, 1988) reported moment-specific stress. The *SUDS* is a questionnaire that measures self-reported perceived stress towards an anxiety provoking or a stressful situation, which was used in one study with children with ASD (Lopata et al., 2008). Two questions were provided with a Visual Analogue Scale ranging from 0 to 100 with 0 referring to ‘no stress at all’ or ‘not feeling good at all’ and 100 referring to ‘the most stress you have ever felt’ or ‘the best I have ever felt’. After scoring one question in reverse order, the scores were averaged to create the SUDS composite score. Good to excellent internal consistency based on the SUDS composite score was reported with Cronbach’s alpha ranging between 0.85 and 0.92, as the questionnaire was used in two study conditions. The items were negatively correlated (*r*=-0.74 to −0.85), confirming measurement of the same construct through opposite scaling. In addition, a mild to moderate relationship was found between a physiological measure (cortisol) and the total score on the self-report.

## Discussion

The purpose of this systematic review was to provide an exhaustive overview of the psychometric properties of self-reported measures regarding stress in individuals with ASD, in addition to a description of the underlying construct they are measuring. In total, seven different questionnaires were used in 20 different study populations of individuals with ASD to measure self-reported stress.

Adults and adolescents with ASD were included in studies using the DASS and the modified version of the SSS to assess the level of stress (Bishop-Fitzpatrick et al., 2017a; Cage et al., 2018; George & Stokes, 2018; Jackson et al., 2018; Maddox & White, 2015; McGillivray & Evert, 2014, 2018; Nah et al., 2018; Pahnke et al., 2014; Park et al., 2019; Zimmerman et al., 2017). In addition, the PSS was only administered in adults with ASD (Bishop-Fitzpatrick et al., 2017a, 2017b, 2018; Hirvikoski & Blomqvist, 2015; Hong et al., 2016; Pahnke et al., 2019; Wijker et al., 2020) despite the presence of modified versions of the PSS, including one for adolescents (van der Ploeg, 2013). Up until now, no studies have used the SSS, DASS and PSS in children (aged below 13 years) with ASD, so information regarding feasibility and other psychometric properties of these questionnaires in this young population is lacking. However, the presence of stress in children and adolescents with ASD was examined in other studies using child-adapted questionnaires, such as the SiC and the CSQ-CA. Lastly, both the SUDS (Lopata et al., 2008) and the self-developed questionnaire by Hillier et al. (2016) were used in only one study with children and adolescents with ASD, respectively.

Evidence concerning the unique contribution of self-reports on internalizing states in individuals with ASD has been mentioned in previous research (Berthoz & Hill, 2005; Keith et al., 2019; Rieffe et al., 2011) and has been supported by the studies included in this review. First, the feasibility of the reported questionnaires in various study populations with ASD was confirmed. Second, the absence of significant correlations between self-reports of adolescents with ASD and informant-reports was demonstrated in the study of Pahnke et al. (2014), using the modified version of the SSS. Therefore, it is hypothesized that the content of subjective stress-reports differs from the content gathered by informant-reports (teachers) due to the adolescents’ difficulties with communicating stress towards their teachers or, alternatively, their difficulties with interpreting their own emotional status. These findings are in line with other studies, indicating poor correlations between self-reports and informant-reports of people with psychiatric symptoms, including ASD (Keith et al., 2019; Miller et al., 2014). In addition, it is more sensible to ask individuals themselves on their internalizing states since the experience of emotions and presence of internalizing symptoms are internal processes to which only they have direct access (Barrett et al., 2007; Lambie & Marcel, 2002). Although unique information can be provided by self-reports of individuals with ASD, informant-reports are more commonly used to gain insight into internalizing states of individuals with ASD (Keith et al., 2019). Thus, a sensitization for using self-report tools regarding stress in individuals with ASD is needed. However, these self-report tools should be well validated and reliable.

The results of this systematic review revealed that, although the psychometric properties of some of the included questionnaires have been assessed thoroughly in various populations, this is not the case for populations with ASD. None of the included studies intended to assess the psychometric properties in the populations with ASD. Therefore, they did not meet the prerequisites for methodological scoring according to a standardized assessment tool for psychometric properties such as the COSMIN-checklist (COnsensus-based Standards for the selection of health Measurement Instruments; Mokkink et al., 2010). However, preliminary evidence for construct validity, more specifically defined as hypothesis testing according to the COSMIN-taxonomy, is available for some questionnaires in this review. Therefore, the hypothesis that individuals with ASD would report higher perceived stress than other populations was used. For adults with ASD, higher total scores on the SSS and PSS-14 were reported in comparison to typical peers in only one study for each questionnaire respectively (Bishop-Fitzpatrick et al., 2017a; Hirvikoski & Blomqvist, 2015). This scarce evidence might point towards construct validity for the PSS-14 and SSS in adults with ASD. Four studies using the DASS-21 and one study using DASS-42 reported higher total and subscale scores for adolescents and adults with ASD as compared to typical peers or norm values (Cage et al., 2018; George & Stokes, 2018; Maddox & White, 2015; McGillivray & Evert, 2018; Nah et al., 2018). Therefore, the hypothesis that individuals with ASD have a higher symptom level of depression, anxiety and stress than the general population might be confirmed, although this is based on scarce evidence and only representative for adults with ASD. Moreover, the discriminative capacity of the DASS-21 was insufficient when comparing individuals with ASD and individuals with other psychiatric symptoms. This is not surprising given the high co-occurring rate of psychiatric problems in individuals with ASD (Mannion & Leader, 2013; Matson & Goldin, 2013). Indeed, the presence of any psychiatric disorder might lead to equal or similar amounts of perceived stress but with different levels of impact on daily functioning, which might not be distinguished by using the DASS-21. Thus, the latter might have a sufficient construct validity for identifying individuals from clinical groups versus individuals in the general population but might be insufficient for the discrimination between different clinical groups, especially when clinical groups with high prevalence of co-occurring disorders such as ASD are included. However, this is in contrast with the findings of Antony et al. (1998), who reported differences in scores between several clinical groups and between clinical and nonclinical groups, providing evidence for discriminant validity of both DASS-versions. In general, additional research is needed to support the construct validity of the previously mentioned questionnaires. Moreover, validation studies of the modified version of the SSS still need to be performed (McGillivray & Evert, 2018; Pahnke et al., 2014).

Preliminary evidence for criterion validity was reported for the SUDS as its scores did correlate with a physiological golden standard for stress measurements (cortisol), albeit with a large variation across the results (Klimes-Dougan et al., 2001; Selye, 1950). The authors hypothesized that, based on their results, the self-ratings of children with ASD on the SUDS might be valid when reporting moderate or greater distress but might be invalid when lower levels of distress are reported (Lopata et al., 2008). Although these results are preliminary, further research might enhance the level of evidence and confirm this hypothesis. However, the ongoing discussion on the possible presence of correlations between physiological and self-reported measures on stress in individuals with ASD should be taken into consideration (Romanczyk & Gillis, 2006). Self-reports on stress might uncover unique information concerning this topic, which cannot be provided or confirmed by physiological data. This could be an alternative explanation for the large variation found in the study of Lopata et al. (2008).

In addition to the preliminary results of validity of these questionnaires, some studies reported values of internal consistency as a preliminary indication of reliability properties (Henson, 2001). These results implied a good to excellent internal consistency of the SSS, DASS, CSQ-CA and SUDS, based on the total scores (see Table 2). However, caution must be taken with the interpretation of the subscale ‘Social/Environmental Interactions’ of the SSS as its internal consistency was only poor. The results of the PSS-10 suggested an almost excellent internal consistency, although this result was based on one study sample only (Bishop-Fitzpatrick et al., 2017a, 2018). Finally, no reports on internal consistency or other reliability measures were found for the SiC and PSS-14 in individuals with ASD.

It is remarkable that only two studies used the DASS-42 in a population with ASD whereas the DASS-21 was used in seven studies. This might be explained by the larger number of items in the long version and the superior psychometric properties of the short version, as reported previously in other populations (Antony et al., 1998; Henry & Crawford, 2005). Furthermore, no information was available on the scaling of the self-developed questionnaire of Hillier et al. (2016), which made it difficult to compare its construct with the other questionnaires included in this review.

A comparison of the questionnaires on item level pointed towards differences between the content of the questionnaires. Only two questionnaires in this review (PSS and SiC) fully covered the concept of perceived stress, according to the definition of Phillips (2013), including items concerning symptoms of stress and the ability to cope with them. The DASS and CSQ-CA also included the description of stress-related symptoms but no items on coping abilities. Finally, using the SSS, SUDS and the developed questionnaire of Hillier et al. (2016), participants are asked to rate the intensity of their stress reaction in contrast to rating the frequency of stress-related symptoms as in the previously mentioned questionnaires. In addition, the SSS consists of very concrete descriptions of situations known to be stress provoking in individuals with ASD, whereas the other questionnaires in this review were not developed for individuals with ASD specifically. Therefore, it is important for researchers and clinicians to take into account which concept they aim to measure with self-reported questionnaires concerning stress as not all questionnaires cover the same aspects. This would result in different outcomes, such as a possible referral when using different questionnaires as a screening measure for the same individual.

It is important to note the differences in the examined timespan across questionnaires and between the studies, which complicates the comparison of the results. Some studies implemented a rather broad timespan to examine symptoms, for instance during the past month or past week, using the DASS or PSS (Bishop-Fitzpatrick et al., 2017a, 2017b, 2018; Cage et al., 2018; Hong et al., 2016; Maddox & White, 2015; McGillivray & Evert, 2014, 2018; Zimmerman et al., 2017). Other studies even included the entire lifespan to gather information concerning stress with the PSS in individuals with ASD (Hirvikoski & Blomqvist, 2015; Pahnke et al., 2019). These timespans might induce recall bias (Althubaiti, 2016), which in turn might be different for individuals with or without ASD as frequently observed in clinical practice. Individuals with ASD tend to focus more on one specific stressor and they usually experience more difficulties with describing stress or mood over a longer period. This different perception of stress over time could cause differences in the response pattern on the questionnaires. Although this fell beyond the scope of the included studies, future researchers should consider this possible confounding factor. Furthermore, using the DASS, current symptom assessment was reported as well. Although this might provide valuable information, the momentary assessment of symptom levels may be strongly influenced by the situations that the individual has encountered in the few hours before the administration of the questionnaire in addition to the individual’s mood that day. Thus, assessing current symptoms reflects only a snapshot of the presence of certain symptoms, which is usually not generalizable throughout the participant’s overall mood status. Therefore, a well-evaluated timespan should be considered when using trait-like questionnaires as mentioned above. In contrast, state-like questionnaires such as the SUDS can cover a short time span due to the momentary character of this assessment regarding an individual’s perceived stress; for instance, to evaluate the immediate effect of a certain stressor.

### Clinical relevance

As previously mentioned, higher levels of perceived stress and difficulties with coping have been reported in children and adults with ASD (Bishop-Fitzpatrick et al., 2015, 2017a; Browning et al., 2009; Groden, 2006; Hirvikoski & Blomqvist, 2015; McGillivray & Evert, 2018). Associations between these higher levels of stress and autistic traits such as difficulties with social communication and the presence of restricted, repetitive behaviors have been found in previous research (Hirvikoski & Blomqvist, 2015). Furthermore, individuals with ASD may feel unaccepted by others, leading to more perceived stress and less social support (Cage et al., 2018). Those with higher intellectual capacities may recognize their own social deficits, leading to an increased vulnerability to stress (George & Stokes, 2018). Furthermore, gender and age-related differences in the level of perceived stress have been reported in individuals with ASD. For instance, females reported higher levels of perceived stress related to social events as well as adults aged 25 years and older who perceived social and environmental factors as increasingly stressful as they got older (McGillivray & Evert, 2018). It is also recognized that heightened levels of perceived stress may further compromise social functioning in adults with ASD and negatively influence their quality of life (Bishop□Fitzpatrick et al., 2015, 2017b; Hirvikoski & Blomqvist, 2015; Hong et al., 2016; Park et al., 2019). Therefore, assessment of perceived stress in individuals with ASD with appropriate measurement tools and subsequent treatment is of high clinical interest. Furthermore, by omitting self-reports, the distinct perspective on internally experienced symptoms by the individual with ASD is missing (Keith et al., 2019). In order to achieve an increased use of self-reports in individuals with ASD, adaptations of current self-report tools might be necessary. In contrast, informant-reports should be used only if adaptations in self-report tools are insufficient to gather knowledge concerning the level of perceived stress in certain individuals with ASD.

### Limitations of the study

Some limitations need to be considered. First, only original studies were included in this review, causing the exclusion of possible interesting studies reported as abstracts or conference papers. However, due to their methodology, insufficient information was available to discuss in this review. Second, no risk of bias assessment was performed on the included studies as their design did not meet the prerequisites to be scored according to the COSMIN-checklist, as previously mentioned (Terwee et al., 2012). This stresses the need for future research to focus on studies determining the psychometric properties of the reported questionnaires. However, a thorough screening strategy was applied in this review in order to account for this lack of risk of bias assessment. Third, given the combined character of the DASS, it could be argued that this questionnaire should have been excluded from the systematic review since it did not focus on the measurement of perceived stress only. However, given the absence of a predefined exclusion criterion for combined questionnaires and the presence of a stress-specific subscale, this questionnaire was eventually included for data extraction and further discussion.

Finally, some features concerning the study samples need to be considered as these might limit the interpretation of the results found in this review. First, most study samples represented a male preponderance, similar to what is typically reported in studies concerning individuals with ASD (Giarelli et al., 2010). However, more woman were included in two studies using the DASS-21 (Cage et al., 2018; George & Stokes, 2018) which was attributed to the format of the data collection by means of a survey (Cage et al., 2018), which might attract more female than male responders (Sax et al., 2003). However, a preponderance of female reports might have an impact on the level of reported stress and/or the consequences related to stress. In a sample of typically developing adults, women reported more daily stress with more conflicts, frustrations, daily demands and chronic problems (Matud, 2004). Therefore, questionnaires with gender-specific norms might give more insight into the experience of stress in individuals with ASD. Second, most studies excluded individuals with ASD and intellectual disability. Therefore, the findings from this review are not generalizable to the general population with ASD, which encompasses individuals with lower intellectual abilities as well. However, the PSS-4 was used in a sample of adults with ASD from which one third was diagnosed with an intellectual disability (Bishop□Fitzpatrick et al., 2017b; Hong et al., 2016). This might be explained by the limited number of questions in this questionnaire, making it more feasible to administer in individuals with lower intellectual abilities, although research to confirm this hypothesis needs to be conducted. Third, in the majority of study populations, the mean age of diagnosis was in the adult range (Cage et al., 2018; Hirvikoski & Blomqvist, 2015). This is not in accordance with common practice where the mean age of diagnosis occurs primarily in childhood or early adolescence due to early detection, screening procedures and the fact that ASD is a neurodevelopmental condition (Elsabbagh et al., 2012; Lai et al., 2014). However, this shift in mean age of diagnosis might be partly explained by the large proportion of females in one of these studies (Cage et al., 2018) for whom a diagnosis might be found later in life in comparison to males with ASD (Giarelli et al., 2010). Finally, in some studies, the participants were not recruited using strict inclusion criteria (Cage et al., 2018), especially in one study where no detailed information regarding diagnosis or diagnostic procedures was provided (George & Stokes, 2018). Furthermore, in the study of Jackson et al. (2018), 20 participants scored below the cut-off criterion of the Autism Quotient (AQ-10) but were still included in the group with ASD as the authors suggested that these participants had false negative scores. All previously mentioned factors are important to consider when interpreting the results of this review since they refer to heterogeneous representations of populations with ASD, as is commonly reported in literature.

### Implications for future research

Despite the presence of clear, clinical relevance of assessing self-reported stress levels and the feasibility of administering such tools in individuals with ASD, evidence of psychometric properties of these self-reports is still scarce. This gap in current research should be addressed by using appropriate study designs in future research. For instance, by including typically developing peers and populations with other clinical disorders than ASD, the hypothesized construct validity as mentioned in this review can further be investigated. Furthermore, the collection of normative and gender-specific data on self-reported measures in individuals with ASD can provide useful insights into screening for stress-related complaints in these individuals (McGillivray & Evert, 2014; Ozsivadjian et al., 2014). In addition, repeated assessments might provide more insight into reliability and responsivity features of the reported questionnaires in this review. Following standardized guidelines, such as the COSMIN-checklist, can increase the homogeneity in future study designs. In addition, the examined timespan should be mentioned to enhance comparability of different study results. Feasibility studies of the SSS, DASS and PSS in children and adolescents with ASD need to be conducted in addition to studies focusing on the psychometric properties in this population. This could be combined with adapting the questions according to the developmental and age-specific situations that this population encounters. Finally, the reliability and quality of current self-reports in individuals with ASD and intellectual disability might be lower due to their limited ability to reflect upon their inner state. However, future researchers should aim to develop adapted versions of self-reports to increase the feasibility of use by simplifying the questions and using more concrete language. In addition, an adapted version of informant-reports, as proposed by Hong et al. (2016), could be used for the assessment of perceived stress in this population. This adapted version inquires information of how the parents think their child would respond to the questions (Sheldrick et al., 2012) instead of typical other-reports, where parents are asked to estimate the perceived stress of their child (Li et al., 2015). Correlations between self-reports and these adapted informant-reports were higher compared to correlations between self-reports and ‘typical’ informant-reports. This argues for the use of adapted informant-reports in order to gather information on a certain topic whenever participants are unable to answer themselves (Hong et al., 2016). However, it should be noted that the questionnaires used were inquiring information on quality of life, for which the adapted informant-reports might be more feasible than for topics related to the experience of stress. In sum, a combination of the previously mentioned adaptations regarding self-reports and informant-reports could enhance the knowledge of self-reported stress in individuals with ASD and intellectual disability even more and should be addressed in future research.

## Conclusion

This review included seven different questionnaires based on 22 studies regarding self-reported stress in individuals with ASD. It is important to keep in mind which concept of stress researchers aim to measure as not all questionnaires encompass the same aspects of perceived stress. Based on the self-report questionnaires found in this review for adults and children with ASD, only the PSS and the SiC respectively cover the concept of perceived stress whereas the other questionnaires reflect upon the frequency or intensity of symptoms of stress. Future research should examine the psychometric properties of the questionnaires included in this review for individuals with ASD, as the current evidence on psychometric properties is too scarce to recommend the use of one of these questionnaires. Finally, it might be necessary to combine this research with the implementation of ASD-specific adaptations of the questions in order to enhance the comprehensibility in this population.

## Supporting information

Electronic Supplementary Material 1

Electronic Supplementary Material 2

## Data Availability

Not applicable

## Conflict of interest

On behalf of all authors, the corresponding author states that there is no conflict of interest.

## Acknowledgements

The authors wish to thank Thomas Vandendriessche, Kristel Paque and Krizia Tuand, the biomedical reference librarians of the KU Leuven Libraries – 2Bergen – learning Centre Désiré Collen (Leuven, Belgium), for their help in conducting the systematic literature search.

## Declarations

### Funding

The research reported was supported by funding from the Marguerite Marie Delacroix Foundation (awarded to AN) and by a postdoctoral fellowship from the Fund for Scientific Research Flanders awarded to KE (12L6916N).

### Conflict of interest

On behalf of all authors, the corresponding author states that there is no conflict of interest.

### CRediT Author Statement

**Anoushka Thoen:** Conceptualization, Methodology, Investigation, Writing – Original Draft, Visualisation. **Jean Steyaert:** Writing – Review & Editing. **Kaat Alaerts:** Writing – Review & Editing. **Kris Evers:** Conceptualization, Writing – Review & Editing. **Tine Van Damme:** Conceptualization, Validation, Writing – Original Draft, Supervision.

## Notes

### Competing Interest Statement

The authors have declared no competing interest.

### Clinical Trial

This study contains a systematic review thus no trial ID is coupled to this study.

### Funding Statement

The research reported was supported by funding from the Marguerite Marie Delacroix Foundation (awarded to AT) and by a postdoctoral fellowship from the Fund for Scientific Research Flanders awarded to KE (12L6916N).

### Author Declarations

This study consists of a systematic review and therefore, is exempt from ethical approval.

## REFERENCES

Althubaiti, A. (2016). Information bias in health research: definition, pitfalls, and adjustment methods. Journal of multidisciplinary healthcare, 9, 211–217.

Antony, M. M., Bieling, P. J., Cox, B. J., Enns, M. W., & Swinson, R. P. (1998). Psychometric properties of the 42-item and 21-item versions of the Depression Anxiety Stress Scales in clinical groups and a community sample. Psychological assessment, 10(2), 176.

American Psychiatric Association. (1980). Diagnostic and statistical manual of mental disorders (3rd ed.).

American Psychiatric Association. (1987). Diagnostic and statistical manual of mental disorders (3rd ed., rev.).

American Psychiatric Association. (1994). Diagnostic and statistical manual of mental disorders (4th ed.).

American Psychiatric Association. (2000). Diagnostic and statistical manual of mental disorders (4th ed., text rev.).

American Psychiatric Association. (2013). Diagnostic and statistical manual of mental disorders (5th ed.)

Baron, M. G., Lipsitt, L. P., & Goodwin, M. S. (2006). Scientific foundations for research and practice. In M. G. Baron, J. Groden, G. Groden & L. P. Lipsitt (Eds.), Stress and coping in autism (pp. 42–67). Oxford University Press.

Barrett, L. F., Mesquita, B., Ochsner, K. N., & Gross, J. J. (2007). The Experience of Emotion. Annual Review of Psychology, 58(1), 373–403.

Barrios, B. A., & Hartmann, D. P. (1988). Fears and anxieties. In E. J. Mash & L. G. Terdal (Eds.), Guilford behavioral assessment series. Behavioral assessment of childhood disorders (pp. 196–262). Guilford Press.

Berthoz, S., & Hill, E. L. (2005). The validity of using self-reports to assess emotion regulation abilities in adults with autism spectrum disorder. European Psychiatry, 20(3), 291–298.

Bishop-Fitzpatrick, L., Mazefsky, C. A., & Eack, S. M. (2018). The combined impact of social support and perceived stress on quality of life in adults with autism spectrum disorder and without intellectual disability. Autism, 22(6), 703–711.

Bishop-Fitzpatrick, L., Minshew, N. J., Mazefsky, C. A., & Eack, S. M. (2017a). Perception of Life as Stressful, Not Biological Response to Stress, is Associated with Greater Social Disability in Adults with Autism Spectrum Disorder. J Autism Dev Disord, 47(1), 1–16.

Bishop□Fitzpatrick, L., Mazefsky, C. A., Minshew, N. J., & Eack, S. M. (2015). The relationship between stress and social functioning in adults with autism spectrum disorder and without intellectual disability. Autism Research, 8(2), 164–173.

Bishop□Fitzpatrick, L., Smith DaWalt, L., Greenberg, J. S., & Mailick, M. R. (2017b). Participation in recreational activities buffers the impact of perceived stress on quality of life in adults with autism spectrum disorder. Autism Research, 10(5), 973–982.

Browning, J., Osborne, L. A., & Reed, P. (2009). Research section: A qualitative comparison of perceived stress and coping in adolescents with and without autistic spectrum disorders as they approach leaving school. British Journal of Special Education, 36(1), 36–43.

Cage, E., Di Monaco, J., & Newell, V. (2018). Experiences of autism acceptance and mental health in autistic adults. Journal of Autism and Developmental Disorders, 48(2), 473–484.

Choque, N. O., Flygare, O., Coco, C., Görling, A., Råde, A., Chen, Q., Lindstedt, K., Berggren, S., Serlachius, E., Jonsson, U., Tammimies,K., Kjellin, L., Bölte, S. (2017). Social skills training for children and adolescents with autism spectrum disorder: a randomized controlled trial. Journal of the American Academy of Child & Adolescent Psychiatry, 56(7), 585–592.

Cohen, S., Kamarck, T., & Mermelstein, R. (1983). A global measure of perceived stress. Journal of Health and Social Behavior, 24(4), 385–396.

Cohen, S. (1988). Perceived stress in a probability sample of the United States. In S. Spacapan & S. Oskamp (Eds.), The Claremont Sympposium on Applied Social Psychology. The Social Psychology of Health (pp. 31–67). Sage Publications, Inc.

De Bruin, E. I., Sieh, D. S., Zijlstra, B. J., & Meijer, A.-M. (2018). Chronic childhood stress: psychometric properties of the chronic stress questionnaire for children and adolescents (CSQ-CA) in three independent samples. Child indicators research, 11(4), 1389–1406.

DuBois, D., Ameis, S. H., Lai, M.-C., Casanova, M. F., & Desarkar, P. (2016). Interoception in autism spectrum disorder: A review. International Journal of Developmental Neuroscience, 52, 104–111.

Elsabbagh, M., Divan, G., Koh, Y.-J., Kim, Y. S., Kauchali, S., Marcín, C., Montiel-Nava, C., Patel, V., Paula, C. S., Wang, C., Yasamy, M. T., & Fombonne, E. (2012). Global Prevalence of Autism and Other Pervasive Developmental Disorders. Autism Research, 5(3), 160–179.

George, R., & Stokes, M. A. (2018). A quantitative analysis of mental health among sexual and gender minority groups in ASD. Journal of Autism and Developmental Disorders, 48(6), 2052–2063.

Giarelli, E., Wiggins, L. D., Rice, C. E., Levy, S. E., Kirby, R. S., Pinto-Martin, J., & Mandell, D. (2010). Sex differences in the evaluation and diagnosis of autism spectrum disorders among children. Disability and Health Journal, 3(2), 107–116.

Groden, J., Baron, M. G., & Groden, G. (2006). Assessment and coping strategies. In M. G. Baron, J. Groden, G. Groden & L. P. Lipsitt (Eds.), Stress and coping in autism (pp. 15–41). Oxford University Press.

Groden, J., Diller, A., Bausman, M., Velicer, W., Norman, G., & Cautela, J. (2001). The development of a stress survey schedule for persons with autism and other developmental disabilities. Journal of Autism and Developmental Disorders, 31(2), 207–217.

Henry, J. D., & Crawford, J. R. (2005). The short-form version of the Depression Anxiety Stress Scales (DASS-21): Construct validity and normative data in a large non-clinical sample. The British Journal of Clinical Psychology, 44, 227–239.

Henson, R. K. (2001). Understanding Internal Consistency Reliability Estimates: A Conceptual Primer on Coefficient Alpha. Measurement and Evaluation in Counseling and Development, 34(3), 177–189.

Hillier, A., Greher, G., Queenan, A., Marshall, S., & Kopec, J. (2016). Music, technology and adolescents with autism spectrum disorders: The effectiveness of the touch screen interface. Music Education Research, 18(3), 269–282.

Hirvikoski, T., & Blomqvist, M. (2015). High self-perceived stress and poor coping in intellectually able adults with autism spectrum disorder. Autism, 19(6), 752–757.

Hong, J., Bishop-Fitzpatrick, L., Smith, L. E., Greenberg, J. S., & Mailick, M. R. (2016). Factors associated with subjective quality of life of adults with autism spectrum disorder: Self-report versus maternal reports. Journal of Autism and Developmental Disorders, 46(4), 1368–1378.

Jackson, S. L., Hart, L., Brown, J. T., & Volkmar, F. R. (2018). Brief report: Self-reported academic, social, and mental health experiences of post-secondary students with autism spectrum disorder. Journal of Autism and Developmental Disorders, 48(3), 643–650.

Jonsson, U., Olsson, N. C., Coco, C., Görling, A., Flygare, O., Råde, A., Chen, Q., Berggren, S., Tammimies, K., & Bölte, S. (2019). Long-term social skills group training for children and adolescents with autism spectrum disorder: a randomized controlled trial. European child & adolescent psychiatry, 28(2), 189–201.

Keith, J. M., Jamieson, J. P., & Bennetto, L. (2019). The importance of adolescent self-report in autism spectrum disorder: Integration of questionnaire and autonomic measures. Journal of abnormal child psychology, 47(4), 741–754.

Klimes-Dougan, B., Hastings, P. D., Granger, D. A., Usher, B. A., & Zahn-Waxler, C. (2001). Adrenocortical activity in at-risk and normally developing adolescents: individual differences in salivary cortisol basal levels, diurnal variation, and responses to social challenges. Dev Psychopathol, 13(3), 695–719.

Lai, M.-C., Lombardo, M. V., & Baron-Cohen, S. (2014). Autism. The Lancet, 383(9920), 896–910.

Lambie, J. A., & Marcel, A. J. (2002). Consciousness and the varieties of emotion experience: a theoretical framework. Psychol Rev, 109(2), 219–259.

Li, M., Harris, I., & Lu, Z. K. (2015). Differences in proxy-reported and patient-reported outcomes: assessing health and functional status among medicare beneficiaries. BMC Med Res Methodol, 15, 62.

Lopata, C., Volker, M. A., Putnam, S. K., Thomeer, M. L., & Nida, R. E. (2008). Effect of social familiarity on salivary cortisol and self-reports of social anxiety and stress in children with high functioning autism spectrum disorders. Journal of Autism and Developmental Disorders, 38(10), 1866–1877.

Lovibond, P. F., & Lovibond, S. H. (1995a). The structure of negative emotional states: Comparison of the Depression Anxiety Stress Scales (DASS) with the Beck Depression and Anxiety Inventories. Behaviour research and therapy, 33(3), 335–343.

Lovibond, S. H., & Lovibond, P. F. (1995b). Manual for the Depression, Anxiety and Stress Scales (2nd ed.). Sydney: Psychology Foundation.

Maddox, B. B., & White, S. W. (2015). Comorbid social anxiety disorder in adults with autism spectrum disorder. Journal of Autism and Developmental Disorders, 45(12), 3949–3960.

Mannion, A., & Leader, G. (2013). Comorbidity in autism spectrum disorder: A literature review. Research in Autism Spectrum Disorders, 7(12), 1595–1616.

Matson, J. L., & Goldin, R. L. (2013). Comorbidity and autism: Trends, topics and future directions. Research in Autism Spectrum Disorders, 7(10), 1228–1233.

Matud, M. P. (2004). Gender differences in stress and coping styles. Personality and Individual Differences, 37(7), 1401–1415.

McGillivray, J., & Evert, H. (2014). Group cognitive behavioural therapy program shows potential in reducing symptoms of depression and stress among young people with ASD. Journal of Autism and Developmental Disorders, 44(8), 2041–2051.

McGillivray, J. A., & Evert, H. T. (2018). Exploring the Effect of Gender and Age on Stress and Emotional Distress in Adults With Autism Spectrum Disorder. Focus on Autism and Other Developmental Disabilities, 33(1), 55–64.

Mendelson, T. (2013). Stress, Emotional. In M. D. Gellman & J. R. Turner (Eds.), Encyclopedia of Behavioral Medicine (pp. 1906–1908). Springer New York.

Miller, L. D., Martinez, Y. J., Shumka, E., & Baker, H. (2014). Multiple Informant Agreement of Child, Parent, and Teacher Ratings of Child Anxiety within Community Samples. The Canadian Journal of Psychiatry, 59(1), 34–39.

Mokkink, L. B., Terwee, C. B., Knol, D. L., Stratford, P. W., Alonso, J., Patrick, D. L., Bouter, L. M., & de Vet, H. C. W. (2010). The COSMIN checklist for evaluating the methodological quality of studies on measurement properties: A clarification of its content. BMC Medical Research Methodology, 10(1), 22.

Nah, Y.-H., Brewer, N., Young, R. L., & Flower, R. (2018). Brief report: screening adults with autism spectrum disorder for anxiety and depression. Journal of Autism and Developmental Disorders, 48(5), 1841–1846.

Osika, W., Friberg, P., & Wahrborg, P. (2007). A new short self-rating questionnaire to assess stress in children. International journal of behavioral medicine, 14(2), 108–117.

Ozsivadjian, A., Hibberd, C., & Hollocks, M. J. (2014). Brief report: The use of self-report measures in young people with autism spectrum disorder to access symptoms of anxiety, depression and negative thoughts. Journal of Autism and Developmental Disorders, 44(4), 969–974.

Pahnke, J., Hirvikoski, T., Bjureberg, J., Bölte, S., Jokinen, J., Bohman, B., & Lundgren, T. (2019). Acceptance and commitment therapy for autistic adults: an open pilot study in a psychiatric outpatient context. Journal of Contextual Behavioral Science, 13, 34–41.

Pahnke, J., Lundgren, T., Hursti, T., & Hirvikoski, T. (2014). Outcomes of an acceptance and commitment therapy-based skills training group for students with high-functioning autism spectrum disorder: a quasi-experimental pilot study. Autism, 18(8), 953–964.

Park, S. H., Song, Y. J. C., Demetriou, E. A., Pepper, K. L., Norton, A., Thomas, E. E., Hickie, I. B., Hermens, D. F., Glozier, N., & Guastella, A. J. (2019). Disability, functioning, and quality of life among treatment-seeking young autistic adults and its relation to depression, anxiety, and stress. Autism, 23(7), 1675–1686.

Phillips, A. C. (2013). Perceived Stress. In M. D. Gellman & J. R. Turner (Eds.), Encyclopedia of Behavioral Medicine (pp. 1453–1454). Springer New York.

Porges, S. W., Macellaio, M., Stanfill, S. D., McCue, K., Lewis, G. F., Harden, E. R., Handelman, M., Denver, J., Bazhenova, O. V., & Heilman, K. J. (2013). Respiratory sinus arrhythmia and auditory processing in autism: Modifiable deficits of an integrated social engagement system? International Journal of Psychophysiology, 88(3), 261–270.

Ridderinkhof, A., de Bruin, E. I., Blom, R., & Bögels, S. M. (2018). Mindfulness-based program for children with autism spectrum disorder and their parents: direct and long-term improvements. Mindfulness, 9(3), 773–791.

Rieffe, C., Oosterveld, P., Terwogt, M. M., Mootz, S., van Leeuwen, E., & Stockmann, L. (2011). Emotion regulation and internalizing symptoms in children with autism spectrum disorders. Autism, 15(6), 655–670.

Romanczyk, R. G., & Gillis, J. M. (2006). Autism and the physiology of stress and anxiety. In M. G. Baron, J. Groden, G. Groden, & L. P. Lipsitt (Eds.), Stress and coping in autism (pp. 183–204). Oxford University Press.

Sax, L. J., Gilmartin, S. K., & Bryant, A. N. (2003). Assessing Response Rates and Nonresponse Bias in Web and Paper Surveys. Research in Higher Education, 44(4), 409–432.

Selye, H. (1950). The physiology and pathology of exposure to stress. Acta, Inc.

Sheldrick, R. C., Neger, E. N., Shipman, D., & Perrin, E. C. (2012). Quality of life of adolescents with autism spectrum disorders: concordance among adolescents’ self-reports, parents’ reports, and parents’ proxy reports. Quality of Life Research, 21(1), 53–57.

Slavich, G. M. (2016). Life Stress and Health: A Review of Conceptual Issues and Recent Findings. Teaching of psychology (Columbia, Mo.), 43(4), 346–355.

Stallknecht, S. E., Strandberg-Larsen, K., Hestbæk, L., & Andersen, A.-M. N. (2017). Spinal pain and co-occurrence with stress and general well-being among young adolescents: a study within the Danish National Birth Cohort. European Journal of Pediatrics, 176(6), 807–814.

Terwee, C. B., Mokkink, L. B., Knol, D. L., Ostelo, R. W., Bouter, L. M., & de Vet, H. C. (2012). Rating the methodological quality in systematic reviews of studies on measurement properties: a scoring system for the COSMIN checklist. Qual Life Res, 21(4), 651–657.

The Oxford Review Encyclopaedia of Terms. (2019). The difference between a state and a trait. Retrieved November 27, 2020, from https://www.oxford-review.com/oxford-review-encyclopaedia-terms/the-difference-between-an-state-and-a-trait/

van der Ploeg, J. (2013). Stress bij kinderen. Bohn Stafleu van Loghum.

World Health Organization. (2016). International statistical classification of diseases and related health problems (10th ed.). https://icd.who.int/browse10/2016/en

World Health Organisation. (2019). International statistical classification of diseases and related health problems (11thed.). https://icd.who.int/

Wijker, C., Leontjevas, R., Spek, A., & Enders-Slegers, M.-J. (2020). Effects of dog assisted therapy for adults with autism spectrum disorder: an exploratory randomized controlled trial. Journal of Autism and Developmental Disorders, 50(6), 2153–2163.

Zimmerman, D., Ownsworth, T., O’Donovan, A., Roberts, J., & Gullo, M. J. (2017). Associations between executive functions and mental health outcomes for adults with autism spectrum disorder. Psychiatry Research, 253, 360–363.

