## Supplementary material for "Self-Reported Stress and Questionnaires in People With Autism Spectrum Disorder: a Systematic Review": Electronic Supplementary Material 2

Review Journal of Autism and Developmental Disorders

Anoushka Thoen, Jean Steyaert, Kaat Alaerts, Kris Evers, Tine Van Damme

Correspondence to Anoushka Thoen, Department of Rehabilitation Sciences, Research Group for Adapted Physical Activity and Psychomotor Rehabilitation, KU Leuven, Herestraat 49 – box 1510, 3000 Leuven, Belgium.

**Search strategy per database**

**Embase**

Concept 1:

‘stress’/exp OR ‘stress*’:ti,ab,kw OR ‘distress’:ti,ab,kw OR ‘mental tension’:ti,ab,kw OR ‘psychological tension’:ti,ab,kw OR ‘psychic tension’:ti,ab,kw OR ‘mental suffering’:ti,ab,kw

Concept 2:

‘autism spectrum disorder’:ti,ab,kw OR ‘Autism’/exp OR ‘Asperger*’:ti,ab,kw OR ‘autistic disorder*’:ti,ab,kw OR ‘high functioning autism’:ti,ab,kw OR ‘autism’:ti,ab,kw OR ‘pervasive child development disorder*’:ti,ab,kw OR ‘pervasive child developmental disorder*’:ti,ab,kw OR ‘pervasive development disorder*’:ti,ab,kw OR ‘pervasive developmental disorder*’:ti,ab,kw OR ‘autistic spectrum disorder*’:ti,ab,kw OR ‘PDD’:ti,ab,kw OR ‘PDD NOS’:ti,ab,kw

**Pubmed**

Concept 1:

"Stress, Psychological"[Mesh] OR stress*[tiab] OR Distress[tiab] OR mental-tension[tiab] OR psychological-tension[tiab] OR psychic-tension[tiab] OR mental-suffering[tiab]

Concept 2:

autism-spectrum-disorder[tiab] OR “Child Development Disorders, Pervasive"[Mesh] OR Autism[tiab] OR Asperger-syndrome[tiab] OR autistic-disorder*[tiab] OR Asperger’s-disease*[tiab] OR Aspergers-disease*[tiab] OR Asperger-disease*[tiab] OR Asperger-disorder*[tiab] OR Asperger’s-disorder*[tiab] OR Aspergers-disorder*[tiab] OR Asperger’s-syndrome*[tiab] OR Aspergers-Syndrome*[tiab] OR Asperger-syndrome*[tiab] OR high-functioning-autism[tiab] OR autism[tiab] OR pervasive-child-development-disorder*[tiab] OR pervasive-child-developmental-disorder*[tiab] OR pervasive-development-disorder*[tiab] OR pervasive-developmental-disorder*[tiab] OR autistic spectrum disorder*[tiab] OR PDD[tiab] OR PDD-NOS[tiab]

**Web of Science**

Concept 1:

"Stress, Psychological" OR “stress*” OR “Distress” OR “mental tension” OR “psychological tension” OR “psychic tension” OR “mental suffering”

Concept 2:

“autism spectrum disorder” OR “Child Development Disorders, Pervasive" OR “Autism” OR “Asperger syndrome” OR “autistic disorder*” OR “Asperger’s disease*” OR “Aspergers disease*” OR “Asperger disease*” OR “Asperger disorder*” OR “Asperger’s disorder*” OR “Aspergers disorder*” OR “Asperger’s syndrome*” OR “Aspergers Syndrome*” OR “Asperger syndrome*” OR “high functioning autism” OR “autism” OR “pervasive child development disorder*” OR “pervasive child developmental disorder*” OR “pervasive development disorder*” OR “pervasive developmental disorder*” OR “autistic spectrum disorder*” OR “PDD” OR “PDD NOS”

**Cochrane library**

#1: [mh “Stress, Psychological”]

#2: (“stress*” OR “Distress” OR “mental tension” OR “psychological tension” OR “psychic tension” OR “mental suffering”):ti,ab,kw

#3: #1 OR #2

#4: [mh “Child Development Disorders, Pervasive”]

#5: (“autism spectrum disorder” OR “Autism” OR “Asperger syndrome” OR “autistic NEXT disorder*” OR “Asperger’s NEXT disease*” OR “Aspergers NEXT disease*” OR “Asperger NEXT disease*” OR “Asperger NEXT disorder*” OR “Asperger’s NEXT disorder*” OR “Aspergers NEXT disorder*” OR “Asperger’s NEXT syndrome*” OR “Aspergers NEXT Syndrome*” OR “Asperger NEXT syndrome*” OR “high functioning autism” OR “autism” OR “pervasive NEXT child NEXT development NEXT disorder*” OR “pervasive NEXT child NEXT developmental NEXT disorder*” OR “pervasive NEXT development NEXT disorder*” OR “pervasive NEXT developmental NEXT disorder*” OR “autistic NEXT spectrum NEXT disorder*” OR “PDD” OR “PDD-NOS”):ti,ab,kw

#6: #4 OR #5

#7: #3 AND #6
